# Detecting space-time clusters of COVID-19 in Brazil: mortality, inequality, socioeconomic vulnerability, and the relative risk of the disease in Brazilian municipalities

**DOI:** 10.1101/2020.06.14.20131102

**Authors:** M. R Martines, R.V Ferreira, R. H. Toppa, L. M. Assunção, M.R. Desjardins, E.M. Delmelle

**Affiliations:** Department of Geography, Tourism and Humanities, Federal University of São Carlos, Sorocaba, SP, Brazil - Research Group: Center for Studies in Landscape Ecology and Conservation; Department of Geography, Federal University of Triângulo Mineiro, Uberaba Campus, State of Minas Gerais, Brazil - Research Group: Center for Studies in Landscape Ecology and Conservation; Department of Environmental Sciences, Federal University of São Carlos, Sorocaba, SP, Brazil - Research Group: Center for Studies in Landscape Ecology and Conservation; Faculty of Law, State University of Minas Gerais, Ituiutaba Campus, Brazil; Department of Epidemiology, Spatial Science for Public Health Center, Johns Hopkins Bloomberg School of Public Health, Baltimore, MD, USA, 21205; Department of Geography and Earth Sciences, Center for Applied Geographic Information Science, University of North Carolina at Charlotte, Charlotte, NC, USA, 28223

**Keywords:** Relative risk, Space-time statistics, COVID-19, Geographic Information Systems, Disease surveillance

## Abstract

The first case of COVID-19 in South America occurred in Brazil on February 25th, 2020. By June 7th, 2020, there were 691,758 confirmed cases, 36,455 confirmed deaths, and a mortality rate of 5.3%. To assist with the establishment of measures for the strategic planning to combat the COVID-19 pandemic in Brazil, we present the first Brazilian geographic study with the aims to examine “active” hand “emerging” space-time clusters of COVID-19. We examine the associations between clusters and mortality rate, vulnerability, and social inequality. We used the prospective space-time scan statistic to detect daily COVID-19 clusters and examine the relative risk from February 25^th^ - June 7^th^, 2020 in 5,570 Brazilian municipalities. We apply a Spearman’s statistic to measure correlation between the relative risk of each cluster and mortality rate, GINI index, and social inequality. We detected 11 emerging space-time clusters of COVID-19 occurring in all Brazilian regions, with seven of them with a relative risk greater than one, and the highest in the Amapá state in the northern region of Brazil. We observed a positive and significant correlation between the relative risk and mortality rate, Brazilian Social Vulnerability Index, and GINI Index. The results can be utilized to improve COVID-19 response and planning in all Brazilian states.

## INTRODUCTION

Over the past 18 years, zoonotic coronavirus transmissions have been a global health concern. During that period, there were two epidemics, first the SARS-CoV, which spread across 30 countries in six continents and resulted in 8,098 cases and 774 deaths (9.5%) (Al-Tawfiq et. al., 2014), while the second was the MERS-CoV, with 1,797 infections between 2012 and 2017 and 687 deaths (38%) across 27 countries, specifically in the Middle East and Korea and affected (Aly et. al., 2017). The third time that a zoonotic coronavirus had crossed species and infect humans is SARS-CoV-2 (Perlman, 2020) – also called coronavirus disease 2019 (COVID-19) –, but it is the first time a coronavirus is a pandemic. The first case of COVID-19 in South America occurred in Brazil on February 25, 2020. The country has a high connection with other countries through airports and ports, especially from cities such as São Paulo, Rio de Janeiro, facilitating the spread of the disease inland and in coastal regions, as well as, to neighboring countries (Rodriguez-Morales et. al, 2020; FIOCRUZ, 2020a).

On June 7^th^, 2020, there were 691,758 confirmed cases, 36,455 confirmed deaths in Brazil, with a lethality rate of 5.3% (Brazil, 2020). However, underreporting of deaths was also expected to bias these numbers (Alonso et al., 2020). It is generally acknowledged that the highest proportion of deaths by COVID-19 occur among the elderly; those with the most severe disease were most likely having a history of hypertension, respiratory disease and cardiovascular disease (Jordan et. al., 2020; Du, et. al., 2020). The risk of death among young adults is smaller than that of older adults, e.g., at most 0.1% – 0.2% (Jordan et al., 2020; Kobayashi et. al., 2020), however severe outcomes and deaths have also been reported among children (Deyà-Martínez et al., 2020; Fabre et al., 2020; Jones et al., 2020). Daniel & Promislow (2020) reported that COVID-19 fatality rates tended to increase exponentially with age, while males tended to have a higher risk of dying at all ages.

COVID-19 has the potential to affect everyone in society; however, the virus affects specific segments of the population very differently, due to their vulnerability (Smith & Judd, 2020). Although Brazil has made significant progress in extending a range of social protection (e.g. universal healthcare), there remains important social inequalities (Landmann-Szwarcwald & Macinko, 2016). The poorest segment of the population is the most vulnerable, especially in the time of a crisis, as it is affected by unemployment, the weakening of social safety nets and access to health services (Ahmed et al., 2020).

Surveillance of COVID-19 is essential to improve response and planning, such as allocating testing and hospital resources, and mitigate the social vulnerability of the population. An effective public health response to the disease requires the ability to monitor and analyze outbreaks under critical space-time conditions. Space-time analytics are particularly attractive to analyze spatial data with a time stamp (Carroll et al. 2014; Delmelle et al. 2011; Delmelle et al. 2014; Jacquez et al., 2005; Levine 2006, Robertson et al., 2010; Rogerson et al., 2008; Paez et al. 2020, Yamada et al. 2009), allowing to estimate the dynamics of infectious diseases. The prospective space-time scan statistic (Kulldorff, 1997) is a widely used cluster detection tool in disease surveillance, which can identify areas that are statistically significant hotspots of disease incidence on the most current time period of the analysis. The statistic determines if the space-time patterns of COVID-19 cases exhibit statistically significant clustering or randomness. Cylindrical scanning windows of different spatial and temporal dimensions are computed to systematically scan the study area and time period for more observed than expected disease cases. The prospective version of the scan statistic is slightly different than the retrospective version (Desjardins et al., 2018; Owusu et al., 2019; Whiteman et al., 2019) because it disregards historical clusters that may have previously existed before the most current day of analysis (Kuldorff 2001).

The prospective space-time scan statistic has been applied to daily case data at the state level in the United States and municipality level in Brazil (Desjardins et al., 2020; Hohl et al., 2020; Ferreira et al. 2020). This approach could improve resource allocation and justify continued social distancing and stay-at-home orders by informing key public health officials about the highest risk areas of COVID-19. The importance of this approach can be extended to analyze the characteristics of the population municipalities within the clusters. As case data are updated, the analysis can be repeated to continuously monitor the evolution of COVID-19 outbreaks (Desjardins et al., 2020). In Brazil, the State Health Secretariats (SHS) update the data daily and make them public, so our approach is well-suited to facilitate daily COVID-19 surveillance in the country. The Ministry of Health reports daily confirmed cases and deaths; while also utilizing a COVID-19 app to disseminate information (de Oliveira et al., 2020). A susceptible, exposed, infected, removed (SEIR) model was applied to several lockdown scenarios (Tarrataca et al. 2020); while Riberio and Bernardes (2020) estimated the number of underreported cases and deaths in Brazil. However, there is currently a lack of spatially explicit studies in Brazil, especially space-time cluster detection of COVID-19 and the associations with vulnerability, mortality rate, and social inequality.

Utilizing a prospective space-time scan statistic, our objective is to detect new emerging clusters of COVID-19 in Brazilian municipalities, considering the data from February 25^th^ to June 7^th^, 2020. When examining these time periods, we intend to compute the evolution of the relative risk of the clusters in different regions and municipalities in Brazil and find associations with mortality rate, vulnerability, and social inequality.

## STUDY AREA

Brazil is comprised of 5,570 municipalities in 26 states. With a population of approximately 210,147,125 people (IBGE, 2019) Brazil is the sixth largest country in population and fifth in landmass, which faces great inequalities and socio-economic disparities. East coast states include approximately 70% of the Brazil’s population. The states of São Paulo and Rio de Janeiro have the highest population density (similar to Europe) while in the states of the Amazon region have densities close to those of Canada and Australia (Somain, 2014).

## DATA AND METHODS

We utilized the SaTScan software for space-time cluster detection of COVID-19 and ArcMap (ESRI, Redlands, CA) for visualization. We compute statistical data in R (version 4.0.1). Data for COVID-19 cases and deaths were retrieved from the Brazil.io project website (Brazil IO, 2020). This project compiles data from the daily COVID-19 case reports by municipality in the 27 units of Brazil and are available in a raw format, which were then tabulated to a format SaTScan could support. The data are from February 25^th^, 2020 to June 7^th^, 2020. In Brazil, 691,758 cases of COVID-19 were confirmed between the dates (Figure 1).

**Figure 1.**
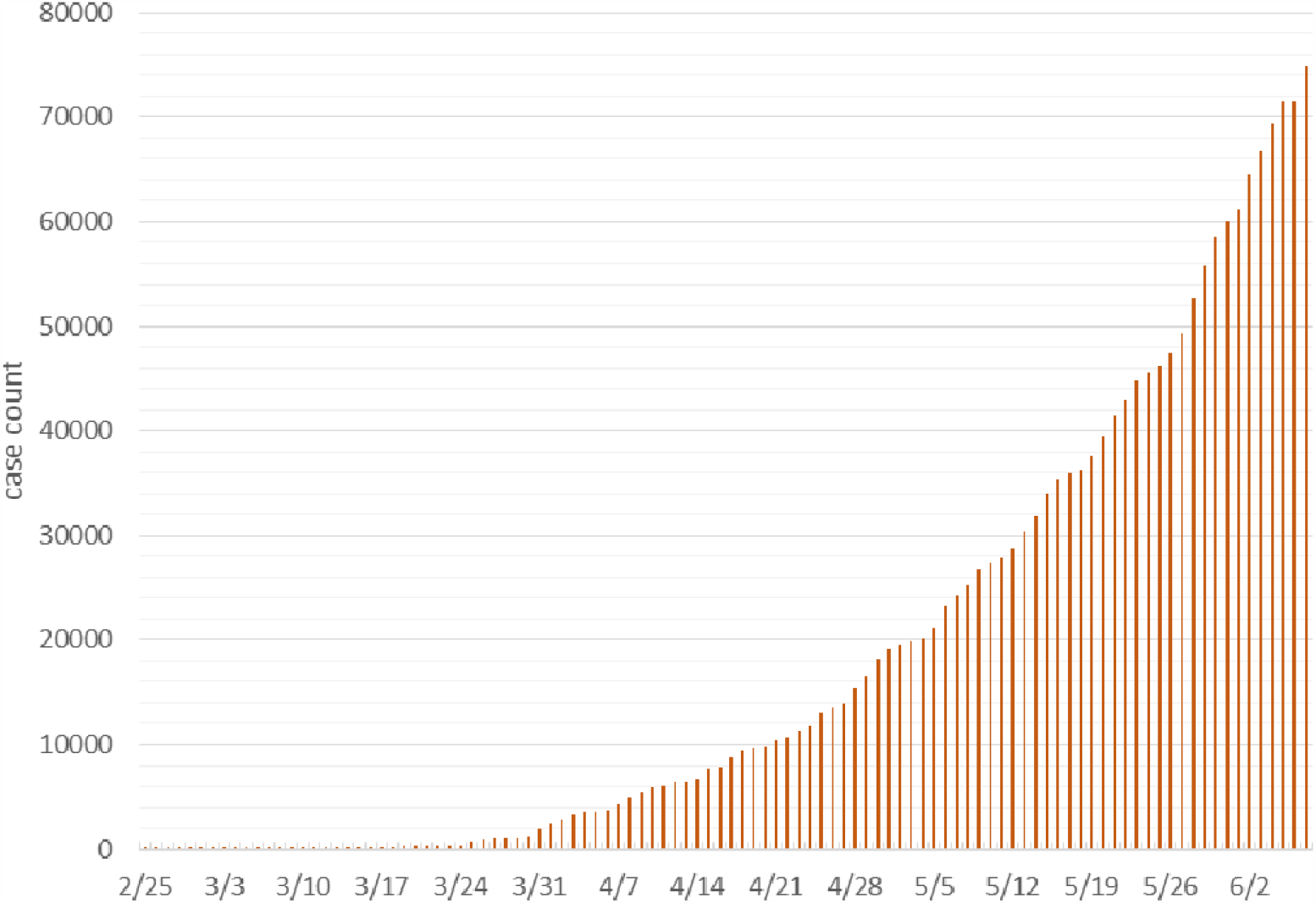
Cumulative number of COVID-19 cases in the State of São Paulo between February 25^th^ and June 07^th^, 2020 (used for the statistical analysis)

In GIS, two layers of geographic data were considered: [1] centroids of the 5,570 municipalities, which was used as a reference of the locations and population for the SaTScan cluster detection; [2] municipality polygons for visualizing clustering and relative risk results via choropleth maps. These layers were retrieved by the Instituto Brazileiro de Geografia e Estatística (IBGE) website; English – The Brazilian Institute of Geography and Statistics.

The detection of active clusters was performed using the prospective Poisson space-time scan statistic method (Kuldorff, 2001). The statistic detects active clusters of disease on the most current day of analysis (Jones et. al, 2006). New data can be added to monitor active and emerging clusters and identify areas that no longer are experiencing excess incidence (e.g. less observed than expected cases). The statistic systematically implements moving cylinders to scan the study area, which are centered on the centroid of the Brazilian municipalities. The base of the cylinder is the spatial scanning window, and the height represents the temporal scanning window; which are both expanded until a maximum threshold is reached. We defined the maximum spatial and temporal search windows to 10% of the population at-risk and 50% of the study period, respectively. Each cluster’s duration was set to a minimum of 2 days and a cluster must contain a minimum of five confirmed cases of COVID-19 (Desjardins et al., 2020).

We utilized a prospective Poisson model to detect space-time clusters that are still occurring or active on June 7^th^, 2020 (Kulldorff, 2001; Kulldorff et al., 1998; Desjardins et al., 2020). We assume that the COVID-19 cases in our study area follow a Poisson distribution under the null hypothesis that states that the model reflects a constant risk. The alternative hypothesis states that the number of observed cases exceed the number of expected cases derived from the null model. The expected cases are estimated by multiplying the population in the cylinder by the total COVID-19 rate in each cylinder.

A maximum likelihood ratio test is implemented to evaluate whether cylinders have an elevated risk for COVID-19. If the cylinder has a likelihood ratio > 1, then it has an elevated risk – (i.e. case rate within the cylinder is greater than the case rate outside of the cylinder). To derive statistical significance, 999 Monte Carlo simulations are computed for each cylinder. We report clusters at the p < 0.05 level, and map the relative risk for each municipality. The relative risk is defined as the estimated risk of COVID-19 within a municipality divided by the risk outside of the municipality.

The results of the clusters were also compared with mortality rate and socioeconomic data. To achieve this purpose, the Gini index (UNDP, 2013) and Brazilian Social Vulnerability Index (SVI) (Brazil, 2015) were used to measure the degree of inequality. The Gini coefficient has been applied in the area of health to measure disparities (Han et. al., 2016) and is based on population income per municipality, and ranges between 0 in the case of perfect equality and 1 in the case of perfect inequality. The SVI is an index that varies between 0 and 1 and summarizes three attributes: urban infrastructure, human capital, and income and labor. The closer to 1, the greater the social vulnerability of a municipality. These dimensions correspond to sets variables that indicates that the standard of living of families is low, indicating non-access and non-observance of social variables. For municipalities with a SVI between 0 and 0.200, this indicates very low social vulnerability; between 0.201 and 0.300 indicates low social vulnerability; between 0.301 and 0.400 indicates middle social vulnerability; between 0.401 and 0.500 indicates high social vulnerability; and between 0.501 and 1 indicates that the municipality has very high social vulnerability (Brazil, 2015). A Spearman’s correlation coefficient was used to measure the correlation (p<0.001) between the relative risk and mortality rate, GINI index, and Brazilian SVI.

## RESULTS

We detected 11 emerging space-time clusters of COVID-19 occurring in all Brazilian regions (p<0.001). Among these clusters, three occurred exclusively in the north and northeast regions (Figure 2).

**Figure 2.**
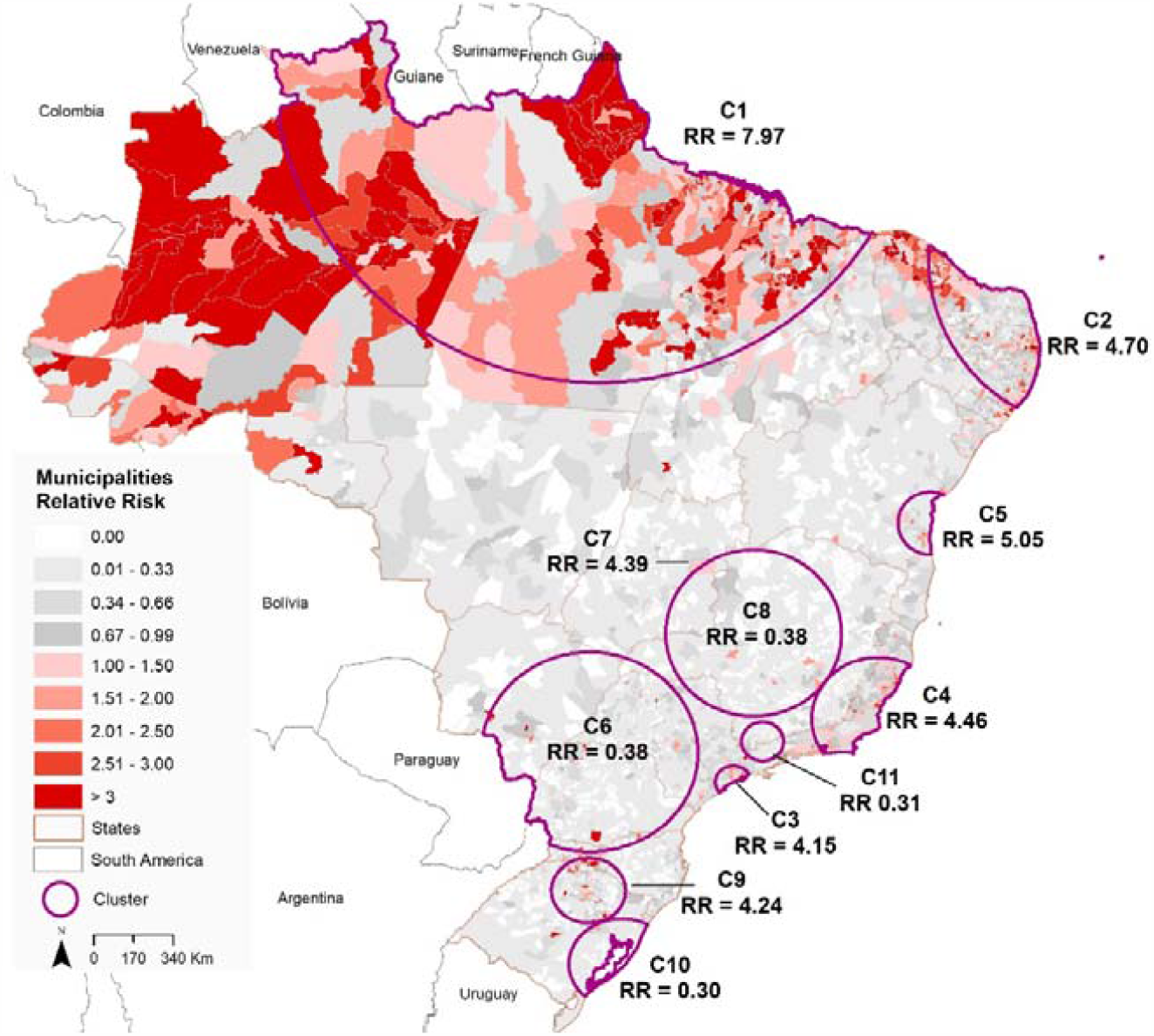
Spatial distribution of emerging space-time clusters of COVID-19 showing the relative risk in Brazil from February 25^th^ to June 07^th^, 2020.

Seven clusters has a relative risk (RR) greater than 1 (i.e. more observed than expected cases) within the period from February 25 ^th^ to June 07 ^th^, 2020 (Figure 3). Cluster 1 (RR = 7.97) is located in the predominantly North region and the state of Tocantins (Center-West region) and includes 466 municipalities, with 293 municipalities showing RR>1. Cluster 2 (RR = 4.7) is found in the Northeast region and includes 584 municipalities, where 180 have a RR>1. Cluster 3 (RR = 4.15) is found in the Southeast region, including São Paulo city and more 34 municipalities, where 15 have a RR>1. Cluster 4 (RR = 4.46) includes 274 municipalities with 48 cities with RR>1 and, also is found in the Southeast region of Brazil covering the states of Minas Gerais, Espírito Santo and Rio de Janeiro. Cluster 5 (RR = 5.05) includes 68 municipalities in the state of Bahia that is in the Northeast region of Brazil, where eight municipalities have a RR>1.

**Figure 3.**
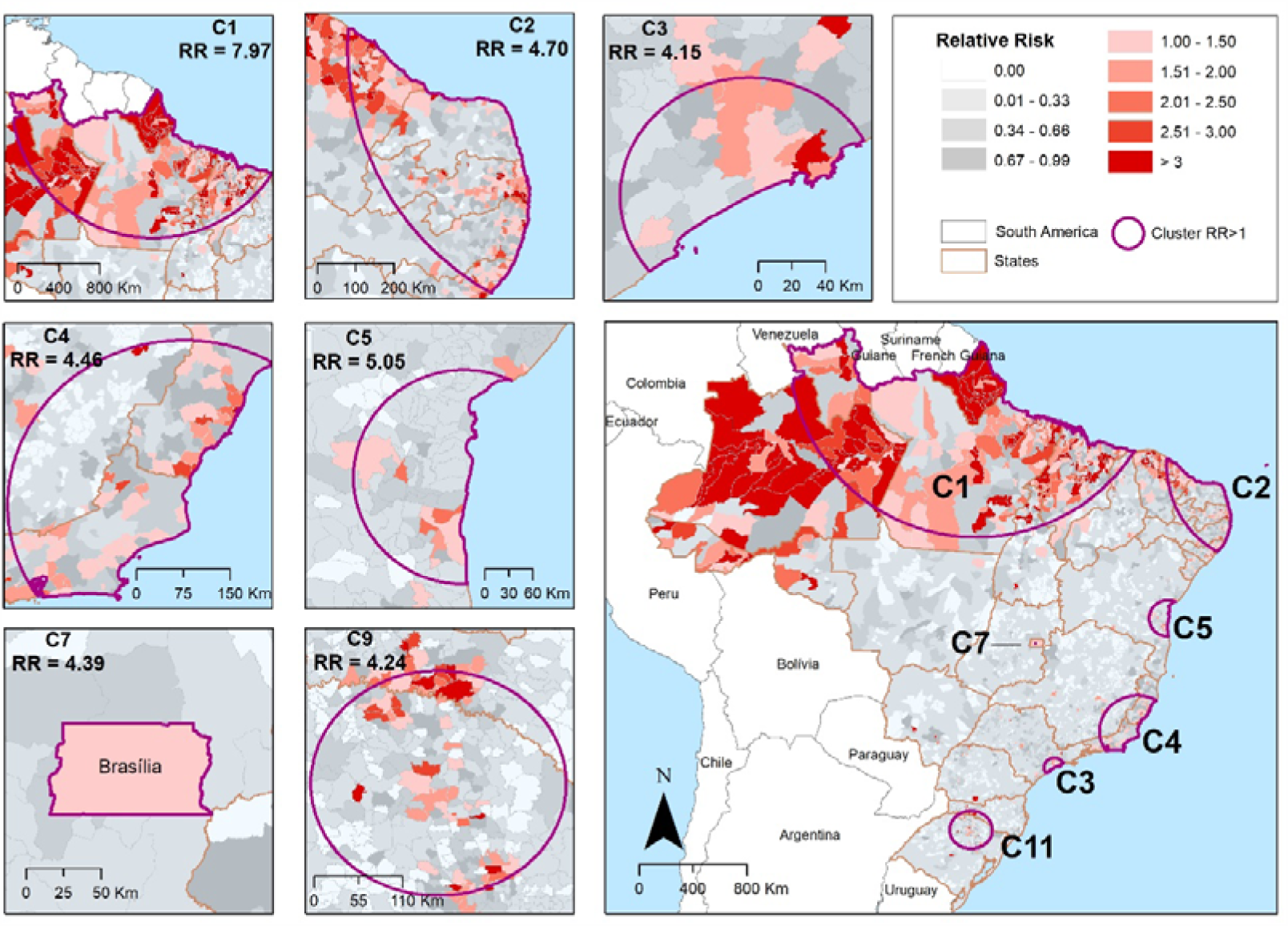
Spatial distribution of emerging space-time clusters of COVID-19 at the municipalities’ level of Brazil from February 25 ^th^ to June 07 ^th^, 2020.

Cluster 7 includes only Brasília, the Capital of Brazil, with a RR of 4.39 located in Center-West region of Brazil. Finally, Cluster 9 (RR = 4.24) includes 230 municipalities located in Santa Catarina and Rio Grande do Sul states, in which 51 municipalities have a RR> 1 (Table 1).

**Table 1.** Emerging space-time clusters of COVID-19 in Brazil from February 25 ^th^ and June 07 ^th^, 2020 (RR = relative risk).

| Cluster | Number of municipalities | Observed | Expected | Cluster RR | Number of Municipalities with RR>1 |
| --- | --- | --- | --- | --- | --- |
| 1 | 466 | 142,789 | 21,929.42 | 7.97 | 293 |
| 2 | 584 | 94,432 | 22,556.14 | 4.7 | 180 |
| 3 | 35 | 81,747 | 21,633.70 | 4.15 | 15 |
| 4 | 274 | 50,151 | 11,909.17 | 4.46 | 48 |
| 5 | 68 | 12,894 | 2,588.76 | 5.05 | 8 |
| 7 | 1 | 9,791 | 2,252.82 | 4.39 | 1 |
| 9 | 230 | 6,222 | 1,476.71 | 4.24 | 51 |

Table 2 shows that the first three municipalities with a RR>1 for each emerging cluster of COVID-19 identified in Brazil from February 25 ^th^ to June 07 ^th^, 2020. We found the highest relative risks in the Amapá state (cluster 1), in the North region of Brazil. The data presented in Table 2 highlighted only two of the Brazilian state capitals (São Paulo, and Vitória) and Brasília, calling attention to the highest relative risks in countryside municipalities and some cities along the shoreline.

**Table 2.** Municipalities with the highest relative risk (RR) for each emerging space-time clusters of COVID-19 in Brazil from February 25^th^ and June 07^th^, 2020.

| *Cluster* | *Region* | *State* | *Municipality* | *Population 2019* | *Observed* | *Expected* | *RR* |
| --- | --- | --- | --- | --- | --- | --- | --- |
| 1 | North | Amapá | Pedra Branca do Amapari | 16,502 | 1,166 | 53.42 | 21.85 |
|  |  |  | Serra do Navio | 5,397 | 289 | 17.47 | 16.54 |
|  |  | Amazonas | Itapiranga | 9,148 | 389 | 29.61 | 13.14 |
| 2 | Northeast | Paraíba | Riachão do Bacamarte | 4,521 | 118 | 14.63 | 8.06 |
|  |  |  | Caaporã | 21,828 | 480 | 70.67 | 6.79 |
|  |  | Ceará | São Gonçalo do Amarante | 48,422 | 993 | 156.77 | 6.34 |
| 3 | Southeast | São Paulo | Santos | 433,311 | 4542 | 1,402.88 | 3.25 |
|  |  |  | São Paulo | 12,252,023 | 74,796 | 39,667.13 | 1.99 |
|  |  |  | Guarujá | 320,459 | 1,829 | 1,037.51 | 1.76 |
| 4 | Southeast | Minas Gerais | Alvarenga | 3,907 | 77 | 12.64 | 6.08 |
|  |  | Espírito Santo | Vitória | 362,097 | 3,508 | 1,172.32 | 3.00 |
|  |  |  | Presidente Kennedy | 11,574 | 112 | 37.47 | 2.98 |
| 5 | Northeast | Bahia | Ipiaú | 45,873 | 337 | 148.51 | 2.26 |
|  |  |  | Itajuípe | 20,491 | 145 | 66.34 | 2.18 |
|  |  |  | Uruçuca | 20,519 | 138 | 66.43 | 2.07 |
| 7 | Center west | Distrito Federal | Brasília | 3,015,268 | 12,864 | 9,762.22 | 1.32 |
| 9 | South | Rio Grande do Sul | Lajeado | 84,014 | 1,389 | 272 | 5.11 |
|  |  | Santa Catarina | Concórdia | 74,641 | 988 | 241.65 | 4.09 |
|  |  |  | Lindóia do Sul | 4,563 | 60 | 14.77 | 4.06 |

We observed a critical situation in the Amapá State, where all the municipalities have a RR>1, and two cases where 50% of the municipalities have a RR>1 (Table 3). We identified this pattern only in Cluster 1 Pará (62.5%) and Maranhão (62.5%). However, it is important to examine the numbers observed in the Amazonas (Cluster 1 – 45.16%), Paraíba (Cluster 2 – 31.39%), Rio de Janeiro (Cluster 4 – 27.17%), Espírito Santo (Cluster 4 – 25.64%), Ceará (Cluster 2 – 23.36%) and Roraima (Cluster 1 – 21.15%) states.

**Table 3.**
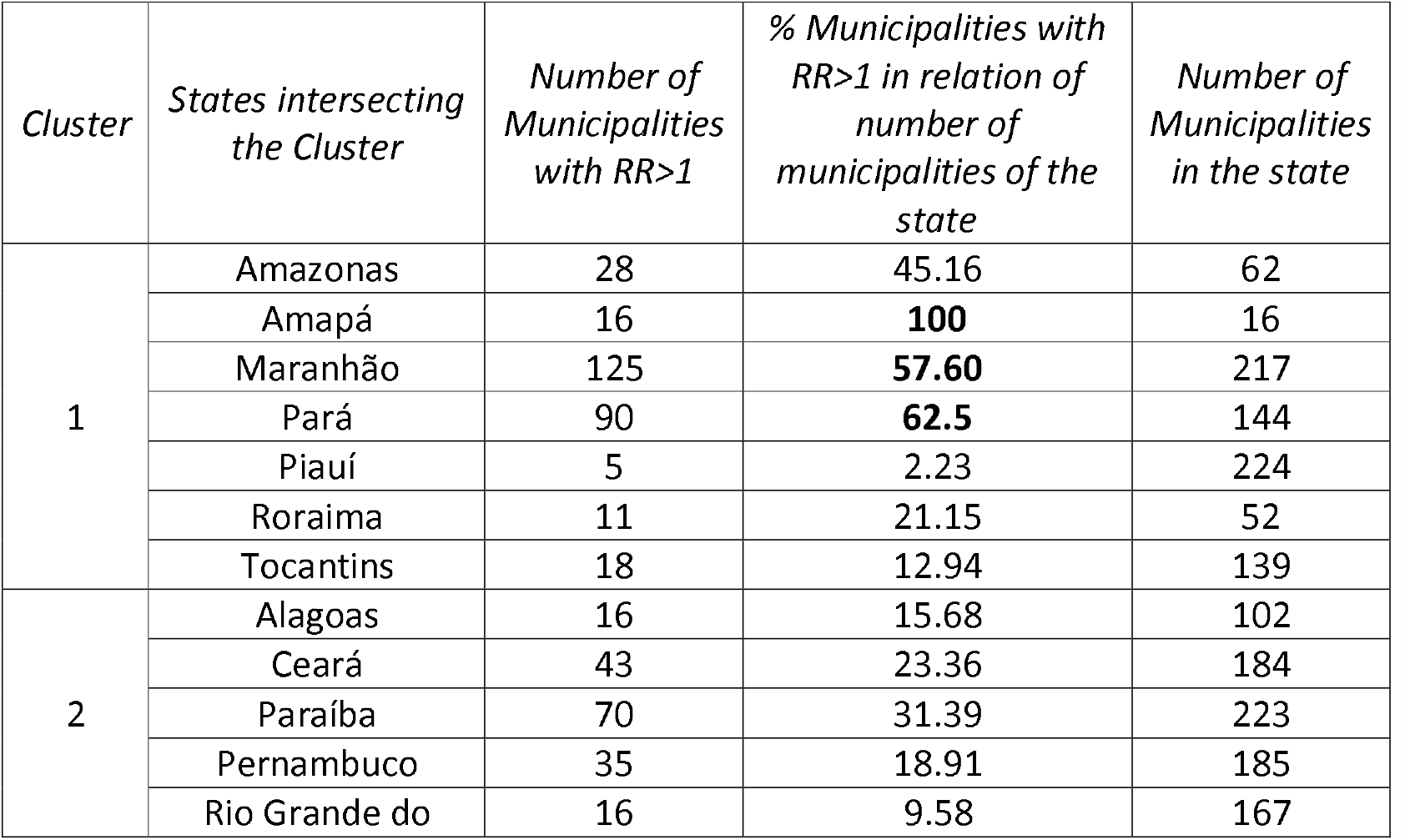
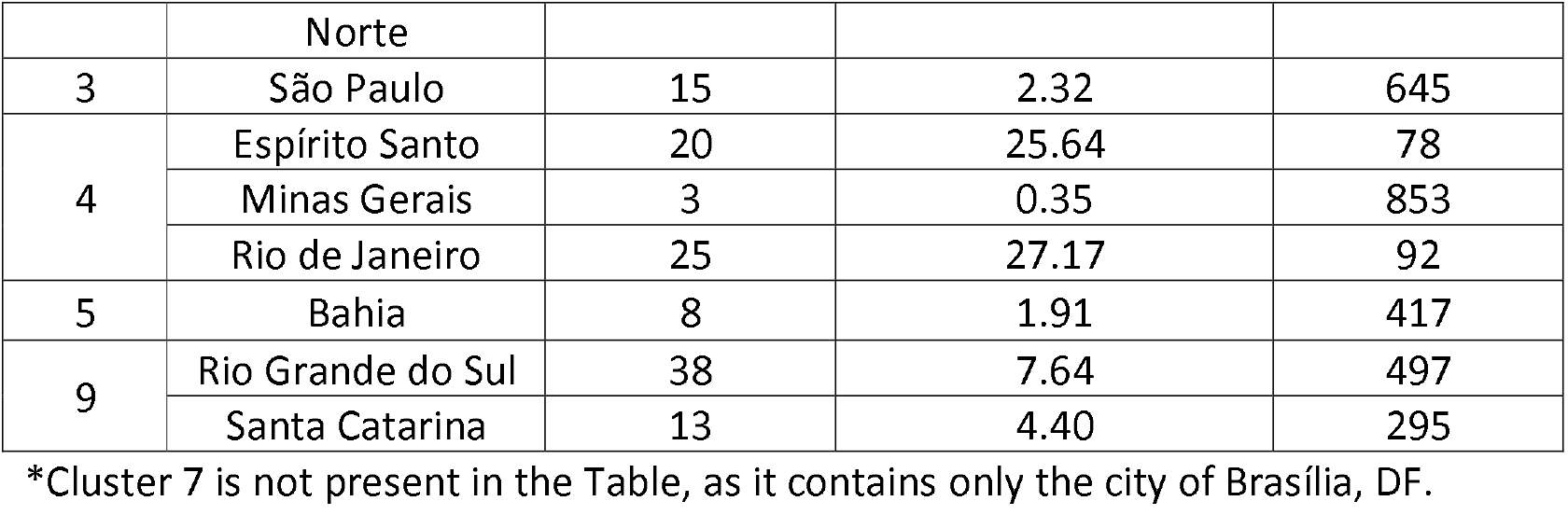
Number of municipalities with relative risk higher than one (RR>1) with the percentage based on the number of municipalities for each state that intersects with the emerging space-time clusters (RR>1) of COVID-19 in Brazil from February 25^th^ to June 07^th^, 2020.

| *Cluster* | *States intersecting the Cluster* | *Number of Municipalities with $RR>1$* | *% Municipalities with $RR>1$ in relation of number of municipalities of the state* | *Number of Municipalities in the state* |
| --- | --- | --- | --- | --- |
| 1 | Amazonas | 28 | 45.16 | 62 |
|  | Amapá | 16 | **100** | 16 |
|  | Maranhão | 125 | **57.60** | 217 |
|  | Pará | 90 | **62.5** | 144 |
|  | Piauí | 5 | 2.23 | 224 |
|  | Roraima | 11 | 21.15 | 52 |
|  | Tocantins | 18 | 12.94 | 139 |
| 2 | Alagoas | 16 | 15.68 | 102 |
|  | Ceará | 43 | 23.36 | 184 |
|  | Paraíba | 70 | 31.39 | 223 |
|  | Pernambuco | 35 | 18.91 | 185 |
|  | Rio Grande do | 16 | 9.58 | 167 |
|  | Norte |  |  |  |
| 3 | São Paulo | 15 | 2.32 | 645 |
| 4 | Espírito Santo | 20 | 25.64 | 78 |
|  | Minas Gerais | 3 | 0.35 | 853 |
|  | Rio de Janeiro | 25 | 27.17 | 92 |
| 5 | Bahia | 8 | 1.91 | 417 |
| 9 | Rio Grande do Sul | 38 | 7.64 | 497 |
|  | Santa Catarina | 13 | 4.40 | 295 |

\*Cluster 7 is not present in the Table, as it contains only the city of Brasília, DF.

Considering all municipalities that are included in a significant emerging space-time cluster of COVID-19 in Brazil, we observed a significant correlation (p<0.001) between the relative risk and mortality rate, Brazilian Social Vulnerability Index (SVI) and GINI index (Figure 4). We found a stronger correlation between the relative risk and mortality rate (Spearman’s rho = 0.605), indicating a tendency that municipalities with higher relative risk are places with more observed deaths. We also observed the same positive correlation between the relative risk Brazilian Social Vulnerability Index (Spearman’s rho = 0.356) and GINI (Spearman’s rho = 0.337) and the, suggesting that municipalities with greater economic inequality and greater social vulnerability may exhibit higher relative risks for COVID-19 cases (Figure 4).

**Figure 4.**
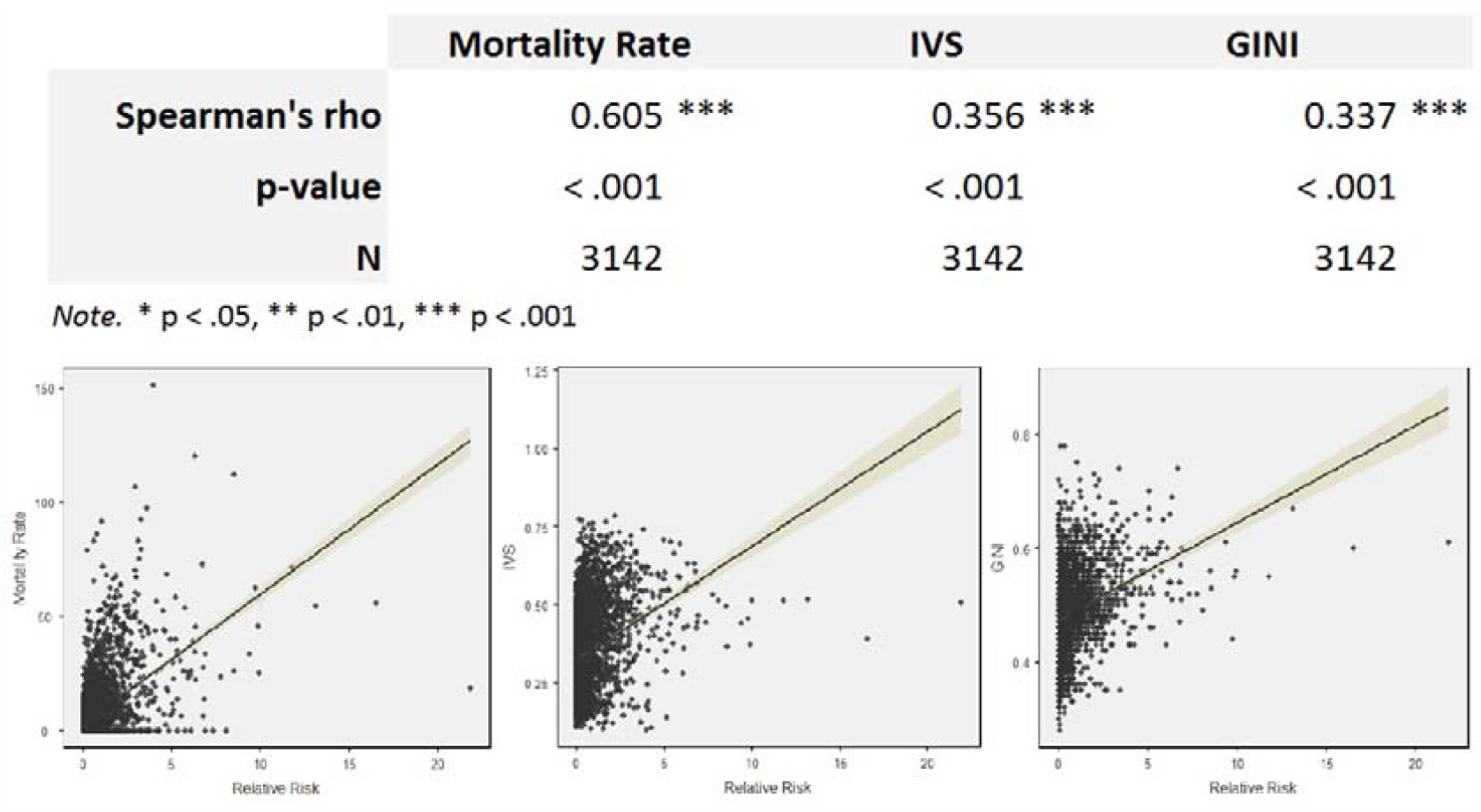
Correlation and Plot with Relative Risk and Mortality Rate, Brazilian Social Vulnerability Index (IVS) and GINI Index.

## DISCUSSION

The results of this study revealed a significant relationship between relative risk and the morality rate, Brazilian SVI and Gini Index. As noted by Figure 4, there is a slight tendency for COVID-19 to manifest itself in clusters with a higher concentration of income, which leads us to infer that this occurs in areas with high social inequality. These findings are in line with a study carried out on the scale of the city of Rio de Janeiro, which focuses on the analysis of the association between the Social Development Index (IDS) and the COVID-19-related Incidence, and Mortality Rate (Cavalcante & Abreu, 2020). Rescaling to the municipality, this relationship begins to appear, although with smaller correlation coefficient.

When bringing to debate the political aspect of the virus, Smith & Judd (2020), consider relevant “to reflect on who is most vulnerable in pandemics”. This question is based on the argument that, despite the fact that COVID-19 can affect the whole of society, its effects will be experienced in different ways, depending on the level of equity that exists in each social reality (Smith & Judd, 2020). As such, it is essential to analyze the pandemic and the policies that emanate from it in the perspective, not only of health, but also of social and economic determinants (Smith & Judd, 2020).

The municipalities with the greatest social vulnerability will ultimately be the locus of incidence and death in Brazil. The high degree of income and access to services are sufficient to suggest that there is “a disproportionate effect of COVID-19 among the most vulnerable in the country” (Pires et al., 2020). In addition, there is a lack of protocols and measures aimed at the social protection of these populations in the atypical context of a pandemic, so when clusters with high risk show a high mortality rate, this can guide decisions for these municipalities.

The slight relationship of active clusters with indices that express inequality in the country may represent the beginning of a problematic scenario, especially so in the most vulnerable municipalities. This may result from the difficulties of enforcing social isolation due to the needs of maintaining employment and income, as well as access to health and basic sanitation (Pires et al., 2020). In the study carried out by Fiocruz (2020) on April 2 ^th^, 2020, the most vulnerable regions of Brazil were identified in the north and northeast regions. In our study, we found that the highest relative risks (RR> 15) were found in the same regions mentioned above, corroborating with their results.

Examining the adherence to social distancing guidelines requires a more detailed analysis, which was beyond the scope of this study. Amazonian communities such as Indians and riverside populations are geographically isolated populations; however, they have been impacted by COVID-19 (ISA, 2020. Therefore, actions need to be taken based on the geographic, social, and cultural differences than those living in urban areas. We present an exploratory study that identifies associations between the relative risk of COVID-19 clusters and social vulnerability, but there are not enough elements to detail demographic particularities of the population, such as sex, age, and ethnicity. An important correlation between clusters with high relative risk and mortality rates, income concentration, and social vulnerability is observed, but the method is sensitive to the scale adopted (Chen et al., 2008) therefore, we hypothesize that the findings will be different and may be more severe regarding relative risk if finer-level data was available. The pandemic is still spreading in Brazil and it is difficult to estimate the speed of transmission along the countryside, where small population municipalities are located. However, our research highlights the regions that are experiencing the highest risk of COVID-19, which is critical for improving public health decision-making. Preventive measures must be strengthened and adhered to, while the only strategy that has proved effective for the control of COVID-19 has been social distancing (de Oliveira et al., 2020).

## CONCLUSION

This research presented an analysis of the dynamics of the expansion of COVID-19 based on the number of daily cases by municipality, with the intent of identifying emerging space-time clusters active in Brazil in the first three months of the pandemic. The results were correlated with data from the socioeconomic condition (mortality rate, GINI and IVS) and a significant positive correlation was identified. We detected seven significant active clusters within Brazil on June 7 ^th^, 2020.

Therefore, this space-time approach to detect emerging clusters will allow decision-makers to identify statistically significant hotspots of COVID-19 cases. States are responsible for coordinating the activities at the regional health level. These regions can use these results to optimize coordination and organization of health care needs, specifically in relation to the poorest populations and those with the highest healthcare demand. Our approach may also allow authorities to pay attention to municipalities that still have little-to-no cases, so they can be prepared to face the burdens of COVID-19. In turn, this can improve the management of resources to the States and Health Regions.

## Data Availability

The COVID-19 data is available from https://Brazil.io/dataset/covid19/

## COMPETING INTERESTS

The authors declare that they have no competing interests.

## Funding

This research did not receive any specific grant from funding agencies in the public, commercial, or not-for-profit sectors.

## Notes

### Competing Interest Statement

The authors have declared no competing interest.

